# Accelerating Tuberculosis Diagnosis in Mozambican Prisons Using Digital Chest X-rays with Computer Aided Detection: Preliminary Results from a Longitudinal, Comprehensive Health Intervention

**DOI:** 10.1101/2024.12.01.24318070

**Authors:** Amadin A. Olotu, Justy Antony Chiramal, Rachel A. Boehm, Aswathy M. Nair, Sanya Chawla, Mário A. Vicente, Dulcidio A. Matusse, Sérgio T. Uate, Amândio S. Munguambe, Edwin J. Prophete, Victoria M. Brown, Cremilde M. Anli, Shibu Vijayan, Ivan R. Calder, Anne C. Spaulding

**Affiliations:** Emory University, Rollins School of Public Health, Center for the Health of Incarcerated Persons (CHIP); National Penitentiary Service of Mozambique (SERNAP); Health through Walls (HtW); Qure ai

**Keywords:** Tuberculosis in prisons, artificial-intelligence, digital-chest x-rays, active TB case-finding, systematic screening for TB, computer-aided detection, tuberculosis preventive treatment, comprehensive health intervention, chronic health conditions, capacity building

## Abstract

**Background:** Mozambique has a high burden of tuberculosis (TB) and in 2021, an estimated 18,000 incident cases nationwide were not diagnosed. Global estimates suggest that 47% of TB disease in prisons is undiagnosed. We implemented an integrated-care-model health intervention to enhance the diagnosis and treatment of TB disease, provide TB preventive treatment (TPT), and identify and treat other undiagnosed health conditions.

**Methods:** Beginning from July 11, 2023, and through the 2nd quarter of 2024 we conducted active case-finding for TB using digital chest x-rays with computer aided detection (DCXR-CAD), in three prisons in Maputo, Mozambique. We conducted clinical assessments for other health conditions and delivered TPT.

**Results:** Of 7912 individuals screened, 264 new cases of TB disease were notified, a TB screening yield of 3.34% and a number needed to screen of 30, and 1346 persons were initiated on TPT. Other conditions were diagnosed and treated including HIV (28), malnutrition (830), and skin conditions (462).

**Conclusions:** Strengthening local infrastructure and implementing DCXR-CAD for screening gave a substantial TB yield in this population. Paying attention simultaneously to preventing TB and addressing other health conditions in this vulnerable population was important.

## 1. Introduction

### 1.1. General Background

Tuberculosis (TB) was the world’s second leading cause of death from a single infectious agent in 2022, causing 1.3 million deaths (1). An estimated 10.6 million persons developed TB disease in 2022, but only 7.5 million people were diagnosed and reported (1). Limited access to healthcare contributes to this gap of 3.1 million cases, that remain undetected by the national health systems (2,3). Key populations who are particularly vulnerable are over-represented among these “Missing Millions”, out of which three-quarters live in 13 of the TB high burden countries (HBC) (2). Concerted global action will be required to achieve the World Health Organization’s (WHO) End TB Strategy’s targets (1,2). One key population especially vulnerable to TB is incarcerated individuals (4), with a TB incidence that is nine-fold higher in prisons than in the general population globally (5), and 11-to 16-fold higher in lower- and middle-income countries (6). Overcrowding, poor ventilation, low healthcare access and malnutrition, found in prisons around the world, contribute to the high burden of TB among carceral populations (6–9). Prisons act as amplifiers for TB, enhancing transmission and reactivation (5,8,10,11). Despite these well-known facts, over 45% of TB cases in prisons go undetected (5). An important dynamic between correctional facilities and the community is a bidirectional flow of TB infections to or from the community (6,11). Mobilizing resources and increasing capacity to systematically screen, detect and treat TB in prisons is therefore critical for any plan to stop TB (6,12) locally, nationally or internationally. This aligns with a core element of the first pillar of the End TB Strategy (13), which aims at achieving early diagnosis and initiation of care for all individuals with a heightened risk for TB. It is equally important to augment access to healthcare for incarcerated individuals, so they may be assessed and treated for other health conditions, since they are disproportionately affected by infectious diseases and non-communicable conditions including chronic physical and mental illnesses (14–17).

### 1.2. Mozambique - Background

Mozambique, occupying the South-Eastern coast of Africa, is one of the ten countries meeting all three WHO measures of TB HBC: it ranks among the 30 countries with the highest incidence of all types of TB cases, of HIV-associated TB cases and multidrug resistant/rifampicin resistant TB cases (1). In 2021, the country had a TB incidence of 363 /100,000 population, and a TB mortality rate of 44/100,000 population (18). Nevertheless, WHO reports that between 2015 and 2022 Mozambique achieved a reduction of 50% in TB deaths (1). In 2021, according to unpublished internal data, among 21,845 incarcerated persons, the National Penitentiary Service of Mozambique (SERNAP) detected 417 cases (1.9%) of TB disease. Like many countries globally (19), the Mozambican correctional system houses a population over its official capacity (20–22), causing overcrowding (20,22), a risk factor for TB and other infectious diseases in prisons (5,7–9). In-prison HIV/TB related deaths have been reported (22). In the past, screening for TB in the prisons of Mozambique was based on residents reporting symptoms at intake or during their stay in prison. Subsequent diagnostic laboratory evaluation utilized a molecular WHO-recommended rapid diagnostic test (mWRD), the GeneXpert MTB/RIF Ultra, which detects the presence of TB disease and rifampicin resistance.

The WHO strongly recommends systematic screening of populations residing in carceral settings for TB (13) and has listed tools and algorithms for enhanced case-finding (13,23). Digital chest X-rays (DCXR) and computer aided detection (CAD) - the use of artificial intelligence (AI) to read and interpret DCXR images, are both tools advocated by the WHO (13,23). Although, they have been evaluated (24–27) and their use in different patient populations reported (28–33), they have not been previously deployed in the penitentiary system of Mozambique. From July 2023, the National Penitentiary Service of Mozambique (SERNAP), with partners - Health Through Walls (HtW), the Center for the Health of Incarcerated Persons (CHIP) at Emory Rollins School of Public Health - commenced a “Health Blitz” in three prisons in Mozambique. The health blitz was the initial phase of a TB REACH-funded longitudinal health intervention and implementation project aimed at addressing TB and other illnesses.

Our objectives in this paper were to: 1) present preliminary results of our intervention of active case-finding for TB using digital chest x-rays with computer aided detection (DCXR-CAD), assessment for other health conditions, and expansion of TB prevention in three prisons in Maputo, Mozambique; 2) examine the proximal effect of the intervention on TB case notifications and notification rates, using an interrupted time-series with comparison group design; and 3) define the role of DCXR-CAD in the clinical diagnosis of TB.

## 2. Materials and Methods

### 2.1. Ethical Considerations

The project protocol was submitted to both the National Committee on Bioethics for Health (CNBS) of Mozambique and the Emory University Institutional Review Board (IRB) for consideration. Clearance to proceed with the “public health intervention” components and approval for the “human subjects research” aspects were obtained from the CNBS and the Emory IRB.

### 2.2. Study Design, Setting and Population

This was a longitudinal public health demonstration project that involved an integrated health intervention delivered to a cohort residing in three prison facilities in Maputo, Mozambique, which was followed longitudinally. An implementation science study which was conducted alongside the demonstration project will be described elsewhere. The correctional facilities were the Maputo City Remand Penitentiary Facility (Remand Prison) with an average daily population (ADP) of 364 male and female residents; the Maputo Provincial Penitentiary Facility (Provincial Prison) with an ADP of 3195 male residents; and the Maximum-Security Special Penitentiary Facility (Maximum Security) with an ADP of 733 male residents. The mean length of stay for residents of the three prisons was: Remand Prison - 2 years [range of 2 months - 2 years], Provincial Prison - 5 years [range of 6 months to 5 years], and Maximum Security - 17 years [range of 8 to 17 years] (Source: SERNAP records). In-prison mortality in 2022 was 29 deaths in the Provincial Prison and 1 death in each of the other two prisons. Of these 31 deaths, 1 was associated with TB alone, and 8 were HIV/TB-related (Source: SERNAP records). Comparison was made with two control prisons that continued delivering the previous standard of care: the Gaza provincial prison (M-Gaza) having an ADP of 514 male residents in Xai-Xai, and the Nampula provincial prison (NRPF) with an ADP of 2158 male residents in Nampula. It is important to note that the population in each of these prisons was not static. Admissions and releases of individuals continued to occur at different rates, but turnover was greatest in the Provincial Prison, with about 23 people admitted and 17 released each weekday.

### 2.3. Inclusion and Exclusion Criteria

All residents who consented to care were included in the health blitz or subsequent screening. Those who were on treatment for TB disease at the time of screening were excluded from the TB screening process but were assessed for other health conditions.

### 2.4. Intervention: Infrastructure and Capacity Building

The “health blitz”, which involved mass screening of the whole population in three prisons and was followed by the screening of entrants through twelve months, was preceded by a planning phase. Prior to the commencement of the health blitz, SERNAP obtained a battery powered mobile digital x-ray machine made by MinXray Inc. (Model TR90BH, Northbrook, Illinois, USA), and qXR (version 3.2, Qure.ai, Mumbai, India) a CAD software. The CAD tool used during the screening process, qXR, is essentially a software that uses AI to read and interpret DCXR. qXR was developed to meet the WHO Target Product Profile (TPP) of at least 90% sensitivity and 70% specificity and has received certification and regulatory clearance for TB screening in humans (13). It has been independently evaluated and was found to meet the WHO TPP (24,26). Its mode of operation is to generate a TB abnormality score that ranges from 0 through 0.50 to 1, depending on the absence or presence of radiological signs of TB in the DCXR. Results of qXR can be viewed on a platform called qTrack, that also collects data. A secondary overlay of the CXR with annotations of abnormalities, which involves highlighting and commenting on anomalous areas (see Figure 1A for a visual representation), and an editable AI report are also generated. Both qXR and qTrack are available in offline mode. SERNAP also acquired an 8-module GeneXpert machine (Cepheid, Sunnyvale CA, USA) and Xpert MTB/RIF Ultra cartridges. Two SERNAP officers were trained in Maputo, over 2 days, by staff of MinXray and Qure.ai, in the use of the X-ray equipment and CAD software. A team comprised of physicians, nurses, other health workers and volunteers were trained for all other aspects of the blitz. Peer educators selected from the population of prison residents were trained to assist in taking weight and height of patients for the calculation of their body mass index. These residents were also trained as patient navigators, to guide fellow prison residents through the different stations during the blitz.

**Figure 1.**
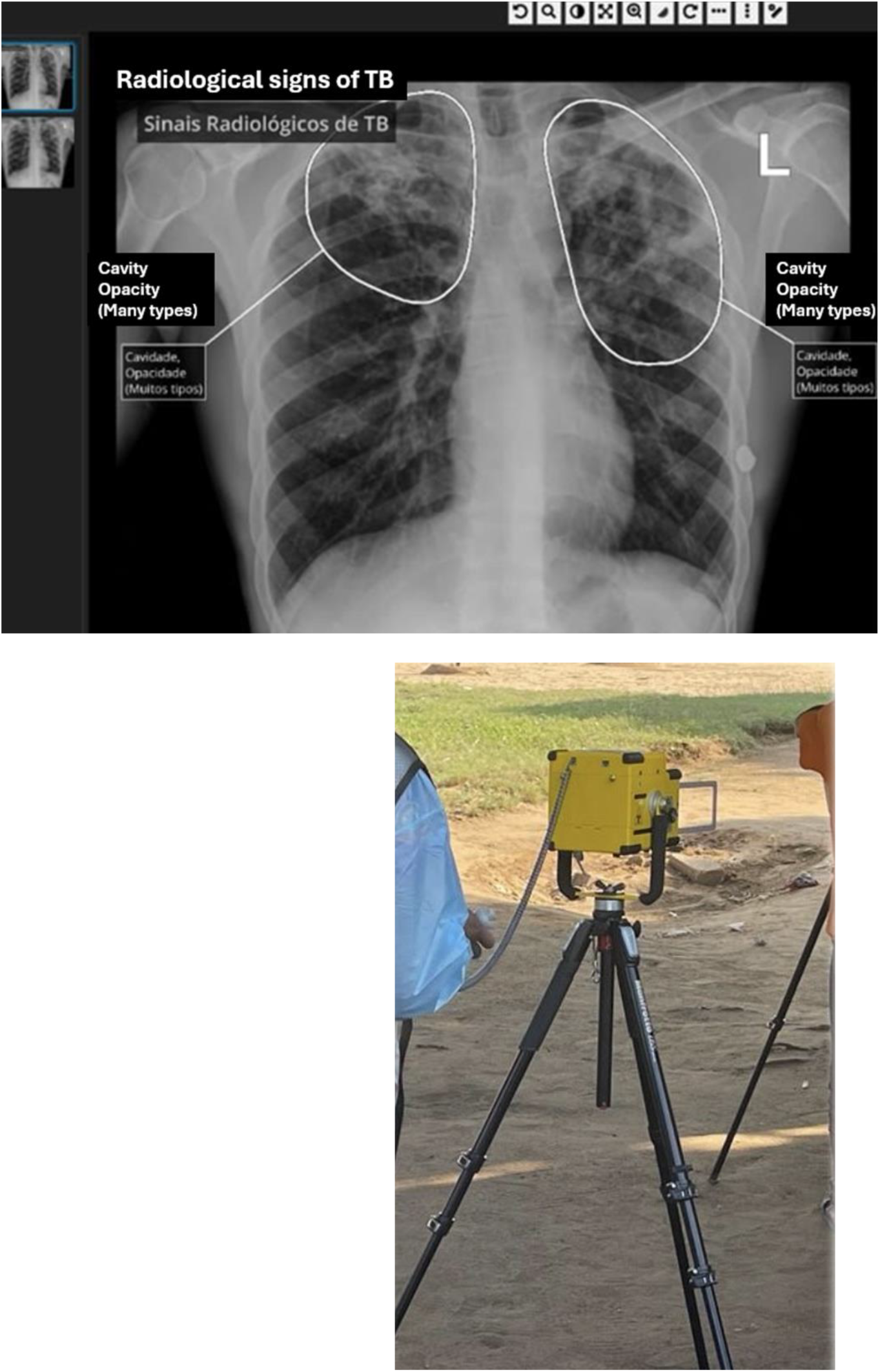
SERNAP officer using mobile digital x-ray machine; & qXR/qTrack user interface. Top: DCXR image on qTrack interface showing qXR interpretation as seen by user abnormalities are highlighted with corresponding comments. Note that text is in Portuguese the official language/language of business in Mozambique, but the English translation has been added. Bottom: SERNAP officer taking a posterior-anterior chest x-ray during blitz.

### 2.5. Intervention: Active TB Case-Finding and Clinical Assessments

Beginning from July 11, 2023, to October 20, 2023 (Remand Prison [July 11 – 14]; Maximum Security [July 17 – 21]; Provincial Prison [July 24 - October 20]), a systematic screening exercise commenced in the three prisons using a parallel screening algorithm. Screening tools were symptom screening (asking about TB symptoms; cough > 2 weeks, hemoptysis, fever, night sweats and unexplained weight loss), and DCXR-CAD; both were done for all residents not on TB treatment. People who had TB symptoms, or with a DCXR-CAD score of 0.50 or greater or with newly positive rapid-HIV test were referred for sputum collection and laboratory diagnostic evaluation with GeneXpert MTB/RIF Ultra. Figure 2 depicts a visual representation of how individuals moved through the different stations. All residents were offered a voluntary opt out HIV rapid test and assessed clinically for other health conditions. Certain individuals who could not produce sputum or in whose sputum *Mycobacterium tuberculosis* (MTB) was not detected, but with a clinical state highly suggestive of TB, were further reviewed by a team of physicians, and received a clinical diagnosis of TB. During the blitz in the Provincial Prison the portable digital x-ray machine broke down and required repair, but the blitz continued and when repair was completed those who had not received x-rays during the five weeks of downtime were recalled and screened with DCXR-CAD. The health screening exercise was universally undertaken by all residents, according to SERNAP officials.

**Figure 2.**
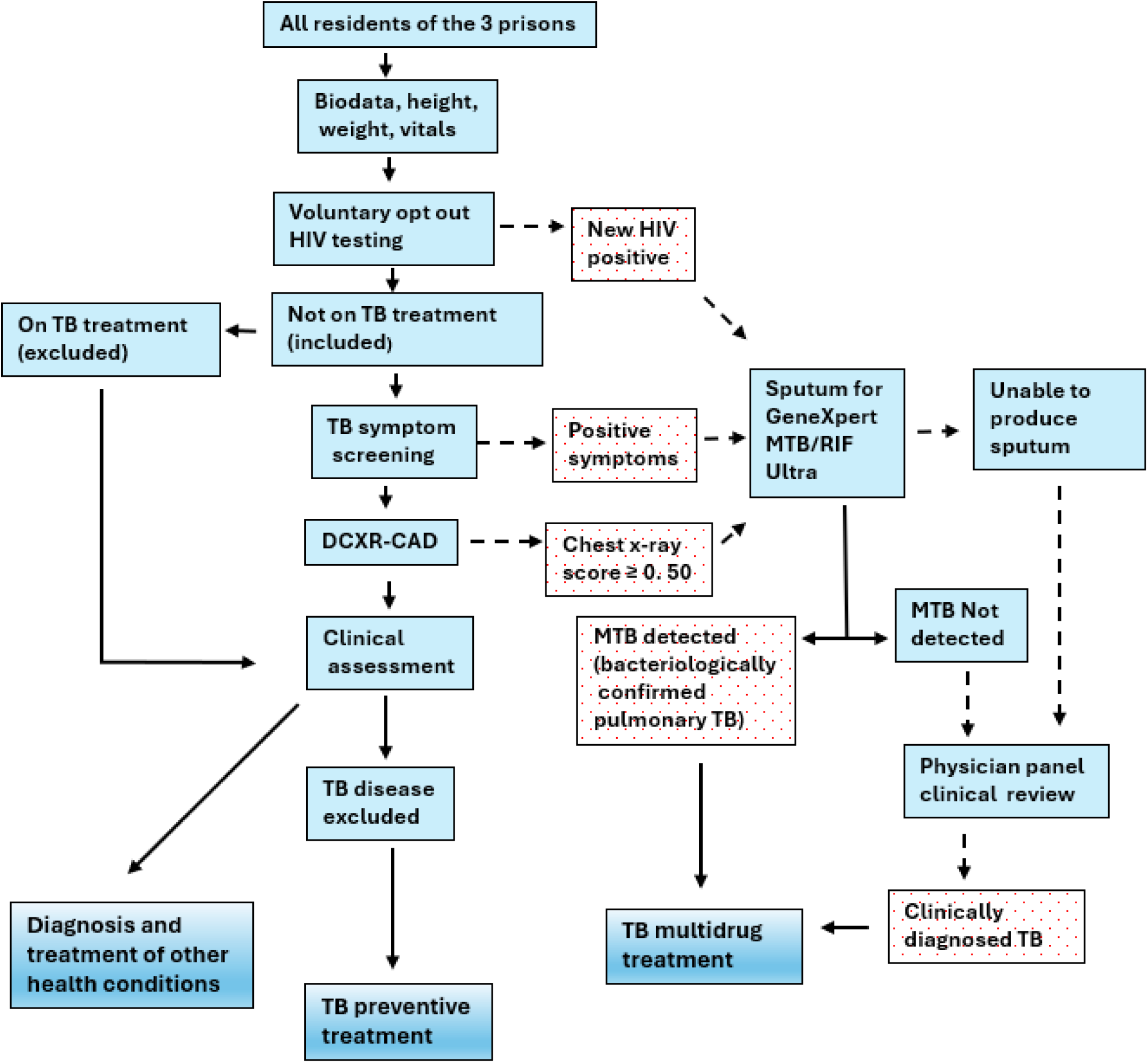
Flowchart of TB Blitz in 3 Mozambican Prisons. Flow chart showing parallel screening algorithm, integrated assessment of other health conditions, pathway to TPT and treatment for TB and other health conditions.

### 2.6. Intervention: Treatment of TB and Other Health Conditions and Provision of TPT

All those with a diagnosis of TB disease were referred to SERNAP healthcare staff for commencement of TB treatment with a standard six-month regimen (isoniazid, rifampicin, pyrazinamide, and ethambutol), by directly observed therapy DOT. Other conditions diagnosed were referred to SERNAP for treatment. A tuberculin skin test (TST) was offered to those at risk, to identify those with tuberculosis infection and guide TB preventive treatment (TPT). Some people, however, did not consent to receiving TSTs. Nevertheless, people in whom TB disease had been excluded, including people with HIV, close contacts of those with TB disease, and those with mental illness, were commenced on three-month TPT using isoniazid and rifapentine (3HP). Peer educators recruited by SERNAP health care staff, were trained in medication monitoring. They were given a special designation of “Chefe de Saúde,” - which translates to English as “the Health Chief” (22). These residents were delegated to assist in conducting DOT for TB treatment and for TPT.

Following the completion of the health blitz, TB screening of newly admitted individuals using DCXR-CAD, in addition to symptom screening, was continued in the three prisons. The mobile digital x-ray machine was rotated between the three prisons on different days of the week to allow the continued use of DCXR-CAD in screening. Treatment for TB disease and TPT using DOT, and treatment for other conditions diagnosed were continued. Those released while on treatment were linked to facilities in the community for continuation of care

### 2.7. Data Collection

Data was recorded manually using clinical and laboratory TB registers routinely used by SERNAP. In addition, healthcare staff used tablets equipped with a mobile data storing application (app) developed by Dimagi Inc. (Cambridge Massachusetts, USA), designated the ‘Health Matters App,’ to collect data. Each tablet was connected to Wi-Fi; data was inputted into the app by staff at each screening station was uploaded to a secure server and routinely synchronized throughout the day. Data was downloaded from the server as comma-separated values (CSV) files. Data management and reconciliation meetings were held by SERNAP to compare and reconcile data collected via the Health Matters App and physical registers and then summarized and reported to TB REACH in quarterly aggregates, which were used for this paper. Other health conditions found during screening and clinical evaluation were recorded as categorical variables and were described using counts and percentages.

### 2.8. Data Analysis

We retrieved data on total TB case notifications counts and bacteriologically positive TB cases for intervention and control prisons aggregated by quarter. Clinically diagnosed TB cases were calculated by subtracting bacteriologically positive TB cases from total TB case notifications. Data was provided for three years preceding the project - beginning from the third quarter of 2020 to the 2nd quarter of 2023. Data from the 3rd and 4th quarters of 2023 reflects the health blitz and continued with screening at intake. The number of people screened in each quarter, when available, were also retrieved for calculation of the case detection/notification rate. The rates were calculated as percentages using the number screened for TB each quarter, as a denominator and total number of TB cases in that quarter as numerator. Quarterly total TB case counts, and calculated TB notification rates, aggregated by prison, were evaluated using an interrupted time-series with comparison group design; a quasi-experimental design that is used for evaluating trends before and after an intervention and incorporates a control group. This was also done for clinically diagnosed TB cases. Data were analyzed using Microsoft Excel 2024 (version 16.89.1) and R (version 4.3.1), and graphs and charts were created using Origin 6.0 software from OriginLab (Northampton, Massachusetts, USA).

## 3. Results

### 3.1 Outcomes

From quarter 3 of 2023 to the 2nd quarter of 2024, 7912 people were systematically screened for TB in all three intervention prisons by symptom screening and DCXR-CAD. Of these 3270 were screened during the health blitz and the rest were new entrants in the post-blitz period. A total of 264 TB case notifications were made for a TB screening yield of 3.34% (264 cases/7912 screenings). The number needed to screen (NNS) was 30 (7912/264). Demographic characteristics of those screened during the blitz and an overview of health conditions other than TB identified during the clinical assessments are shown in Table 1. At the end of the 2nd quarter of 2024 a total of 1,346 people in the three prisons were initiated on TPT.

**Table 1.**
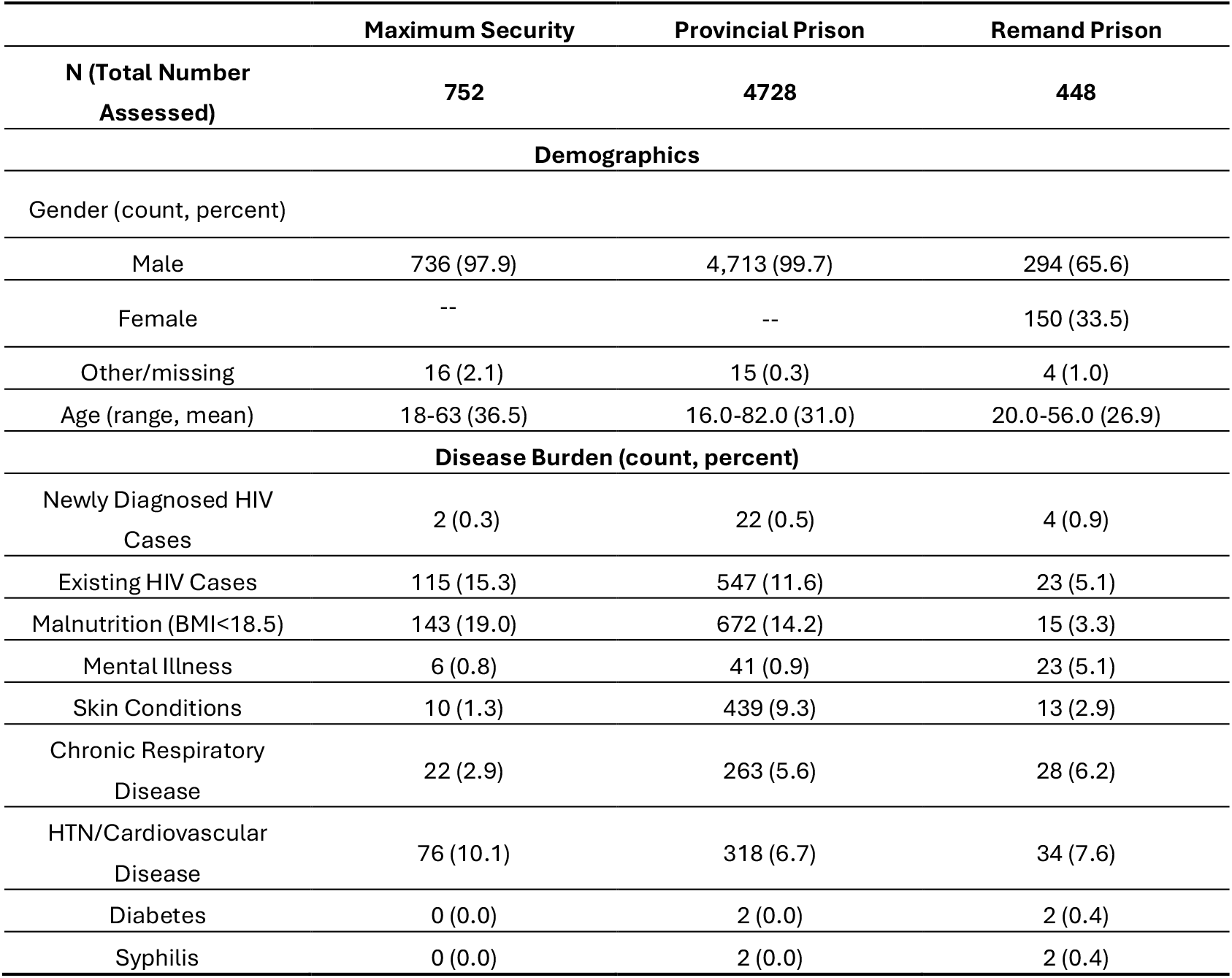
Demographic and disease characterization of intervention prisons in Quarter 3 and Quarter 4 of 2023, Maputo, Mozambique.

|  | Maximum Security | Provincial Prison | Remand Prison |
| --- | --- | --- | --- |
| **N (Total Number Assessed)** | **752** | **4728** | **448** |
| **Demographics** |  |  |  |
| Gender (count, percent) |  |  |  |
| Male | 736 (97.9) | 4,713 (99.7) | 294 (65.6) |
| Female | -- | -- | 150 (33.5) |
| Other/missing | 16 (2.1) | 15 (0.3) | 4 (1.0) |
| Age (range, mean) | 18-63 (36.5) | 16.0-82.0 (31.0) | 20.0-56.0 (26.9) |
| **Disease Burden (count, percent)** |  |  |  |
| Newly Diagnosed HIV Cases | 2 (0.3) | 22 (0.5) | 4 (0.9) |
| Existing HIV Cases | 115 (15.3) | 547 (11.6) | 23 (5.1) |
| Malnutrition (BMI<18.5) | 143 (19.0) | 672 (14.2) | 15 (3.3) |
| Mental Illness | 6 (0.8) | 41 (0.9) | 23 (5.1) |
| Skin Conditions | 10 (1.3) | 439 (9.3) | 13 (2.9) |
| Chronic Respiratory Disease | 22 (2.9) | 263 (5.6) | 28 (6.2) |
| HTN/Cardiovascular Disease | 76 (10.1) | 318 (6.7) | 34 (7.6) |
| Diabetes | 0 (0.0) | 2 (0.0) | 2 (0.4) |
| Syphilis | 0 (0.0) | 2 (0.0) | 2 (0.4) |

### 3.2 Effect of the blitz and post-blitz screening on case notification rates

For each of the three intervention and two control prisons, total case notification counts were available for 16 quarters, 12 before the initiation of the project and four after. The graph for the interrupted time-series with comparison group (or controlled interrupted time-series) design, plotted using total case notification counts for intervention and control prisons is shown in figure 3.

**Figure 3.**
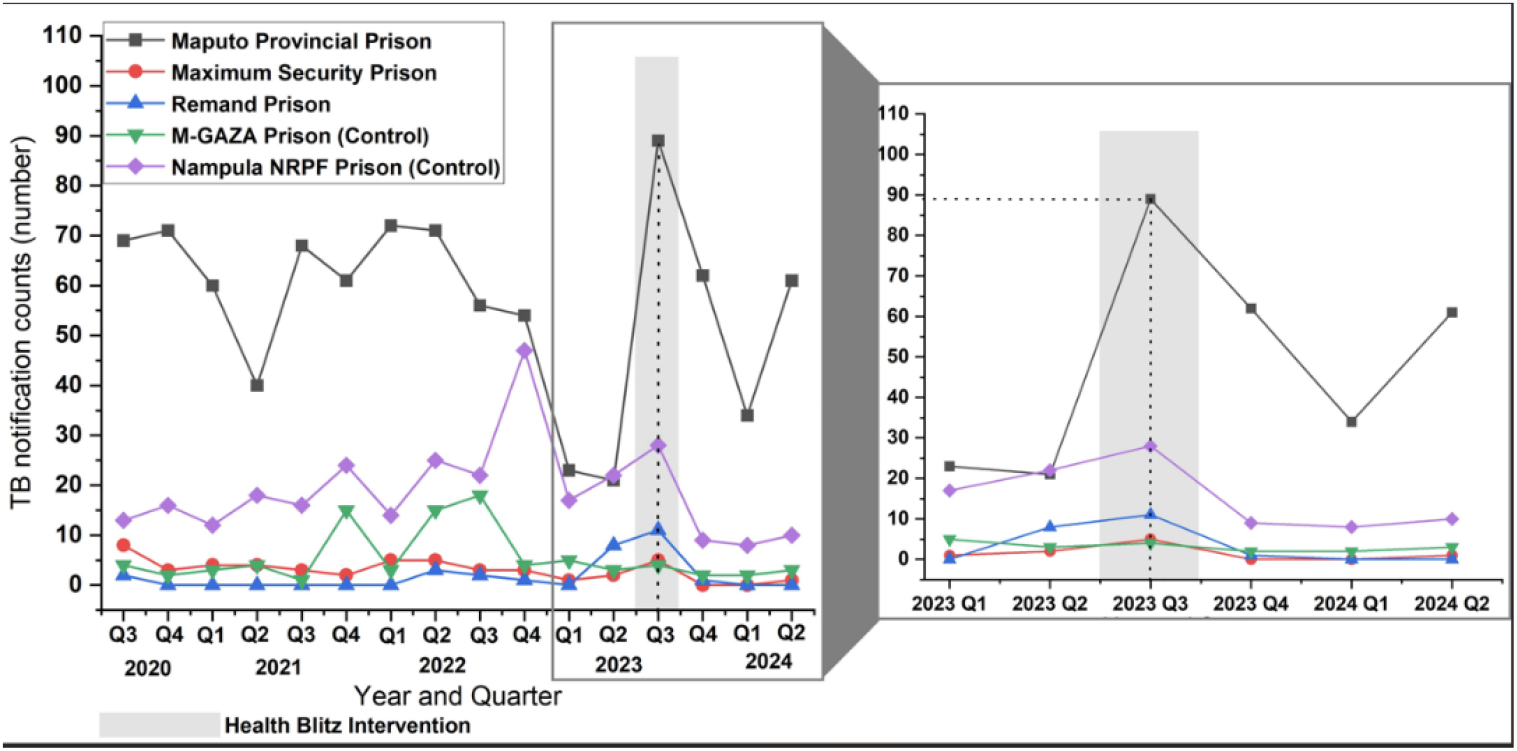
Total TB case notification counts by quarter for each prison. The figure shows all cases of all forms of TB notified in each prison from quarter 3 of 2020 to quarter 2 of 2024. The shaded area represents the health blitz which began during quarter 3 of 2023 and ended early in quarter 4. Screening of new intakes using DCXR-CAD continued in quarters 1 and 2 of 2024.

The trendlines for counts of total TB case notifications in each of the intervention and control prisons, which is shown in Figure 3, display a marked increase in the number of TB notifications in the Maputo Provincial Prison, the largest intervention prison, during the blitz period. An increase is also demonstrated in the other two intervention prisons, the Maximum-Security Prison, and the Remand Prison. This increase is noticeable when the period preceding the blitz is compared with the blitz period for each prison, as counts reach peaks higher than any period before the intervention. The increase during the blitz remains obvious when the intervention prisons are compared with the control prisons that are most similar in population size (Maputo Provincial Prison vs Nampula Prison; Maximum Security vs M-Gaza; Remand Prison vs M-Gaza). No similar upsurge in counts to levels higher than all pre-blitz periods was seen. When the sum of total TB case notifications in the intervention prisons is compared with the sum of total TB case notifications in both control prisons as shown in Figure 4 there is a clear increase in total counts in intervention prisons that is not present in the control prisons during the period of the blitz.

**Figure 4.**
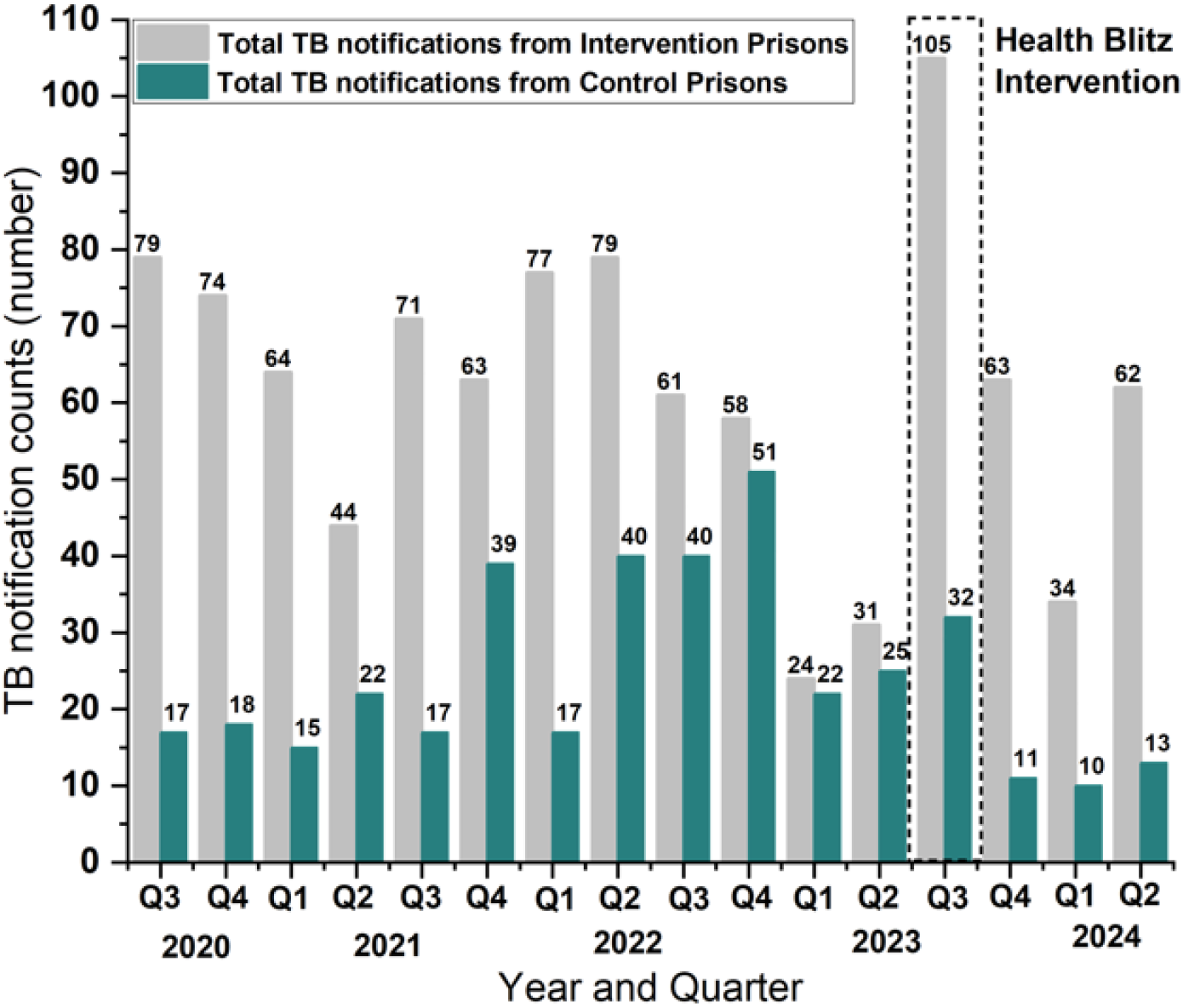
Sum of total TB case notification counts; intervention prisons vs control prisons. Bars correspond to the sum of total TB case notification counts by quarter for all the intervention prisons combined and both control prisons combined. The sum of counts in the intervention prisons during the period of the health blitz markedly exceeds the sum of counts in previous quarters in those same prisons. The control prisons do not show a corresponding increase in that same period.

Numbers screened were not available for control prisons and so case notification rates could not be calculated for them. Case notification rates calculated for the intervention prisons were used to plot a graph (Figure 5) but did not show the same trend of an increase during the period of the blitz, seen with counts. This is understandable given that the numbers screened were relatively lower before the blitz and is particularly highlighted by the most obvious spike which corresponds to quarter 3 of 2022 in the Maximum Security Prison. Out of only 12 people screened in that prison that quarter, 3 cases were detected giving a case notification rate of 25%, the highest in any prison, in any quarter.

**Figure 5.**
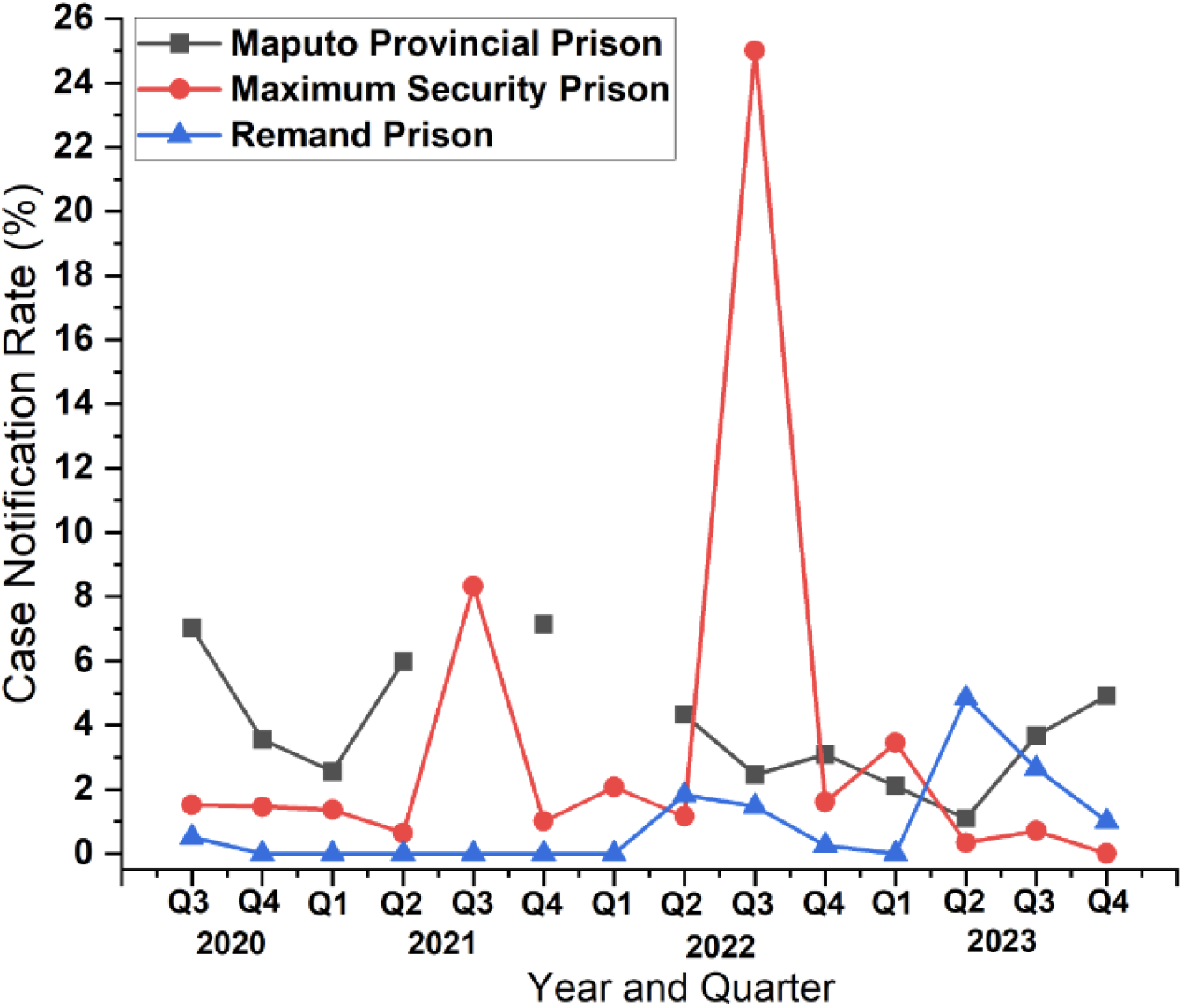
Total TB case notification rates by quarter for intervention prisons. A line graph of case notification rates (cases detected/number screened *100) for all forms of TB notified in the 3 intervention prisons from quarter 3 of 2020 to quarter 4 of 2023. Missing data points in the graph are due to inability to find numbers screened in respective quarters.

### 3.3. Effect of the blitz and post-blitz screening on clinically diagnosed TB cases

There was a considerable increase in the number of TB cases notified that were diagnosed clinically and not bacteriologically confirmed, in the 3rd and 4th quarters of 2023 as well as the 1st and 2nd quarters of 2024. This is shown in Figure 6. However, the increase appears to be limited to the Maputo Provincial Prison, as the same increase is not apparent in the other intervention prisons with smaller populations.

**Figure 6.**
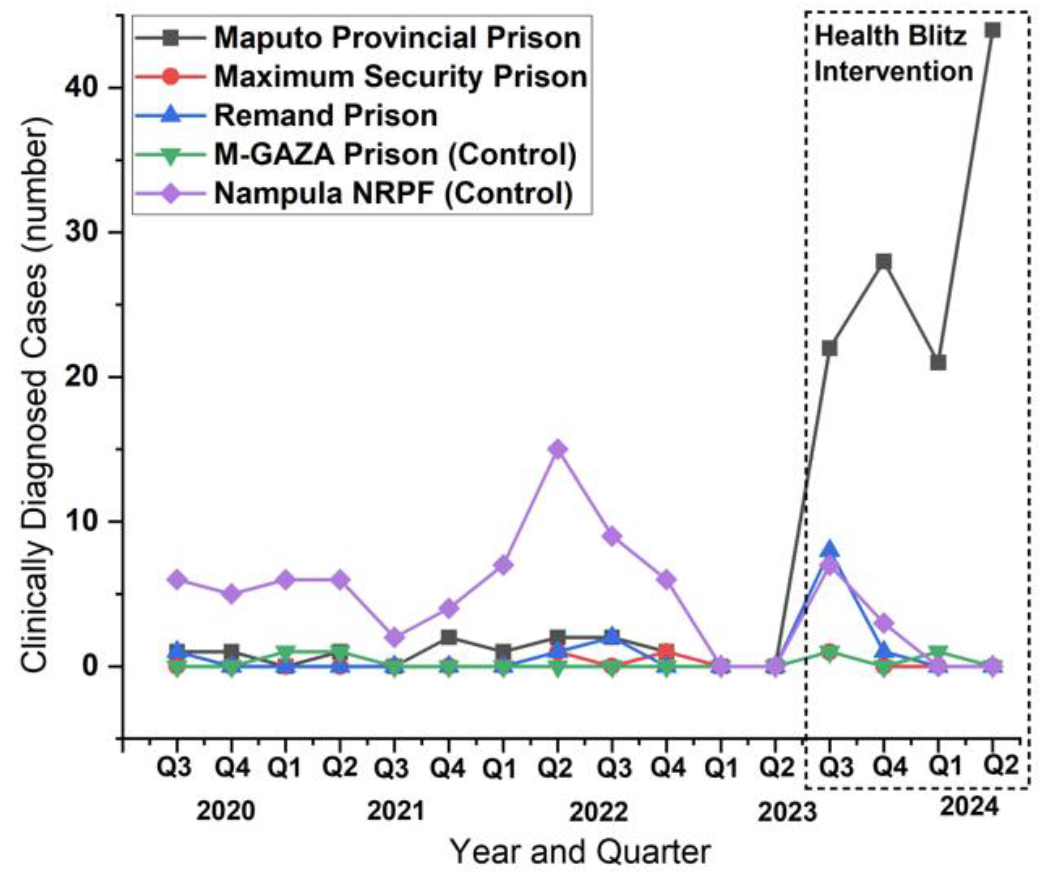
Numbers of clinically diagnosed cases of TB by quarter for each prison. Graph showing a sustained increase in clinically diagnosed TB cases in the Provincial Prison.

## 4. Discussion

After screening 7912 people, a total of 264 TB case notifications made for a TB screening yield of 3.34% and a NNS of 30. The TB yield was considerably higher than that found in other studies in carceral settings in other TB HBC where symptom screening alone was used for active case finding (34–36). It was also higher than where chest X-rays were used in less than 20% of the population (37) or required human interpretation (38). The use of DCXR-CAD in addition to symptoms for screening appears to have contributed to the high yield of TB cases. Studies have found that combining both gives a greater TB yield than using either alone in both prison and community settings (28,39). An alternative approach could have been to offer Xpert MTB/RIF Ultra tests to everyone in the intervention prisons. However, the probable large number of negative results from a higher number of individuals without TB disease would have been very costly in terms of resources, making our approach less cost intensive.

Our evaluation of the immediate effect of the intervention using a controlled interrupted time-series design suggested that the initiation of our project was responsible for the substantial elevation in the total counts of TB notifications that occurred in the intervention prisons during the period of the health blitz in quarter 3 of 2023. This was strengthened by the observation that no similar elevations occurred in the intervention prisons in the years preceding the blitz included in the time-series. Similarly, there were no comparable spikes within that same period of the blitz, in the control prisons. The use of an interrupted time-series design can be strengthened by withdrawing an intervention to see if the observed effect diminishes with the withdrawal. This was not possible with the nature of our intervention and there are ethical issues that make it undesirable to experimentally withdraw health interventions that have been found to be effective or have become the standard of care.

Indeed, many components within the intervention may have contributed to the eventual decline in TB cases. The introduction of TPT, which was happening concurrently with the introduction of the new diagnostics, prevented those in the population with TB exposure from developing TB disease. During the blitz, screening for related conditions like malnutrition and HIV and addressing them could have further decreased new TB cases. Integrating a comprehensive healthcare intervention into a TB screening program could have contributed to the reduction in counts after the blitz.

Our exploration of the role of DCXR-CAD in clinical diagnosis of TB revealed a substantial increase in numbers of clinically diagnosed cases of TB. Remarkably, this increase was sustained into quarters 1 and 2 of 2024. The observation that this upsurge coincided with the initiation of DCXR-CAD in TB screening in the Provincial Prison, and was sustained as its use continued, suggests that it is responsible for the increase. The absence of a similar finding in the pre-intervention period in the Provincial Prison and similar absence in the pre-, intra- and post-blitz periods in the control prisons supports this inference. This might be explained by the understanding that the use of qXR, made available in less than a minute, images, and interpretations of DCXRs with likelihood scores indicating the presence of radiological abnormalities indicative of TB. This provision of radiologic evidence in keeping with TB aided in the clinical reviews of individuals unable to produce sputum or having negative sputum results but with clinical states highly suspicious of TB. The absence of this finding in the other intervention prisons may be attributable to the smaller numbers of TB cases.

We believe adding a mobile digital x-ray machine and CAD software to the existing screening and diagnostic infrastructure owned by SERNAP was instrumental in uncovering numerous, previously hidden TB cases. The improvement of laboratory diagnostic capacity by the addition of an 8 module GeneXpert machine to the SERNAP onsite prison laboratory and procurement of Xpert MTB/RIF Ultra cartridges were necessary to facilitate the progression of the project. The building of human resources and capacity by training SERNAP officers in the use of the newly acquired DCXR-CAD equipment and software tools was critical in achieving project outcomes. Of similar importance was the orientation and training for doctors, nurses, other health workers, volunteers and peer educators who were prison residents. These preparatory steps ensured that it was possible to screen large numbers of individuals within the short time limit of the health blitz using a parallel screening algorithm. Ownership of this new infrastructure by SERNAP made it possible to sustain screening for new intakes into the prison after the blitz period and beyond the project’s duration. This is in line with the WHO’s emphasis on the need to pay attention to planning, human resources, and commodities in preparing for the implementation of screening programs (23). In this project, it was made possible by TB Reach funds, which were used to acquire TB screening tools with enhanced sensitivity.

The cost of procuring the 8 module GeneXpert machine, Xpert MTB/RIF Ultra cartridges, digital x-ray machine and CAD software access for two years was $145,000. Additional unaccounted costs incurred in conducting this intervention, such as labor, if added, would result in a higher figure. We did not conduct a formal cost analysis. Nevertheless, we do recognize that the number of TB infections diagnosed, treated and prevented by this intervention merit consideration. Early diagnosis and treatment of TB in one individual and prevention of progression and transmission by TB treatment and TPT was estimated to result in saving 5,502 US dollars in the Indian health system (40). On the other hand, the indirect economic burden of one person remaining undiagnosed was estimated to be 5,162 US dollars. While this finding may not be generalizable to Mozambican prisons it gives an important perspective on often overlooked economic implications of such TB interventions and the economic costs of undiagnosed TB.

SERNAP’s practice of engaging prison residents as peer educators and as stakeholders and participants in the delivery of their own healthcare increased our staff strength and facilitated the execution of the project. They made assurance of adherence easier even with the sudden increase in the number of people requiring those forms of treatment. Our findings are congruent with published reports that the engagement of residents of prisons and inclusion as peer educators facilitates the successful execution of interventions in carceral settings (41–44).

We described the operationalization of a longitudinal health intervention that initiated systematic screening for TB using DCXR-CAD and symptom screening for active TB case finding, and clinical assessments for other health conditions in 3 Mozambican prisons. We outlined how the treatment for TB and other health conditions diagnosed, and the expansion of TPT delivery were integrated into it. We assessed the effect of our intervention on total TB case notifications and on the clinical diagnosis of TB in these prisons with a quasi-experimental design, a controlled interrupted time-series. To the best of our knowledge, our paper is the first to outline the initiation of a comprehensive health intervention delivered to address TB and other health conditions in a low resource prison setting and evaluate the immediate effect of the intervention on TB case notification. All these aspects of the project addressed the 4 components of the 1st pillar of the End TB strategy (1).

### Strengths and Limitations

Our work was done in prison settings using screening tools that had been previously validated in similar populations. Our findings provide a template for others seeking to initiate similar projects in prisons in TB high burden countries. CAD calibration is ideal when working in a new population to determine the most appropriate threshold, however, we could not do this working in our prison setting, and so we used a threshold of 0.50 to screen and select individuals for sputum evaluation using the Xpert MTB/RIF Ultra assay. Still, qXR, which was the CAD we used, has been found to perform well at that threshold (26). Another limitation was that the mobile digital x-ray machine broke down for a short period during the blitz. Potentially some individuals may have entered and left without receiving a chest x-ray. Once the machine was repaired, the team made a concerted effort to follow up and screen those missed during the down time. Our model of healthcare delivery may not be common in prison systems in other cultures and countries. Employing prison residents to deliver health care may be controversial in carceral settings, where power imbalances can play a significant role. Their participation increased the capacity to move the population through the blitz stations. However, where favoritism is at play, residents delivering healthcare to other residents may lead to undesirable consequences. We explored using statistical tests to compare conditions before and after the intervention and found that not enough measurements were taken for an appropriate analysis using standard methods. This makes our paper more descriptive than quantitative and the absence of a statistical analysis of the extent of change is a limitation.

We did not report on the epidemiological characteristics of all TB cases because our data management system had final reports of TB cases recorded in aggregate, and we were not able to identify epidemiological characteristics by case. We did not assess if there were any associations between the presence or absence of TB disease and factors like age, and length of stay in prison. We hope to explore in future studies the correlation between specific predictors of disease and its development. Multiple confounders could be present, such as immune status, bed proximity to an undiagnosed case of TB, etc., and that future analysis would need to control for these confounders.

## 5. Conclusions

As we approach 2035, we want to retain the hope of achieving the WHO End TB vision of “a world free of TB with zero deaths, disease and suffering due to TB”. The need to focus more resources on the key population in carceral settings is important. Innovative tools and methods found to be effective need to be implemented in prisons. The United Nations Standard Minimum Rules for the Treatment of Prisoners, otherwise known as the Mandela Rules after the former South African president who himself had resided in a prison, espouse the necessity of extending the care available to the public to those residing in prisons. We must adhere to this principle in programs to eliminate TB. DCXR-CAD screening for TB, TPT delivery and assessment and care for other health conditions among incarcerated populations from our experience appear to have a significant role in accelerating progress towards our desirable End TB targets. Nonetheless, there is a need to increase implementation in prisons in TB HBC. We therefore recommend increased attention in prisons in TB HBC with the building of infrastructure to provide services for DCXR-CAD, TPT delivery and assessment and care for other health conditions, in a sustainable model.

## Data Availability

Data Availability Statement: Data used in this paper are available but not in the public domain. They are maintained by the National Penitentiary Service of Mozambique (SERNAP). Requests for data should be made to SERNAP by contacting Dr. Cremilde M. Anli at the following email address

## Author Contributions

Conceptualization, A.A.O, J.C., R.A.B., A.M.N., S.C., M.A.V., D.A.M., S.T.U., A.S.M., E.J.P., V.M.B., C.M.A., S.V., I. R.C. and A.C.S.; methodology, A.A.O, J.C., R.A.B., A.M.N., S.C., M.A.V., D.A.M., S.T.U., A.S.M., E.J.P., V.M.B., C.M.A., S.V., I. R.C. and A.C.S.; validation, D.A.M.; formal analysis, R.A.B., A.M.N., and D.A.M.; investigation, S.T.U., A.S.M., E.J.P., C.M.A. and A.C.S.; resources, I. R.C.; writing—original draft preparation, A.A.O, J.C., R.A.B., A.M.N., S.C., and M.A.V.; writing—review and editing, A.A.O, J.C., R.A.B., A.M.N., S.C., M.A.V., D.A.M., S.T.U., A.S.M., E.J.P., V.M.B., C.M.A., S.V., I. R.C. and A.C.S.; visualization, R.A.B., A.M.N.; supervision, A.C.S; project administration, I. R.C.; funding acquisition, I. R.C. and A.C.S. All authors have read and agreed to the published version of the manuscript.

## Institutional Review Board Statement

The project was conducted according to the guidelines of the Declaration of Helsinki, and ethical approval was obtained from the National Committee on Bioethics for Health of Mozambique (29/CNBS/2023) and the Emory University Institutional Review Board (STUDY00005848).

## Informed Consent Statement

Written informed consent from patients was not required by the institutional review boards on account of all the processes described in this paper being considered a part of a public health intervention and not human subjects’ research.

## Data Availability Statement

Data used in this paper are available but not in the public domain. They are maintained by the National Penitentiary Service of Mozambique (SERNAP). Requests for data should be made to SERNAP by contacting Dr. Cremilde M. Anli at the following email address

We thank the officials and staff of the National Penitentiary Service of Mozambique, volunteers and peer educators (Chefe de Saúde) for their help in executing this project. We thank the residents for their participation. We also thank Qure.ai Research and Development team for the development of the AI device, the Client Success and Business Development team for facilitating the deployment and providing support for any technical difficulties, as well as Dennis Robert from the Clinical Research team for the feedback on data analysis and write up. This work was supported in part by Emory/Georgia Tuberculosis Research Advancement Center (P30 AI168386).

